# Japanese national well-being survey shows that living alone results in lower satisfaction and affects other relationships, while social networking service usage is related to life satisfaction: A cross-sectional study

**DOI:** 10.1101/2022.02.26.22271302

**Authors:** Koichi Miyaki, Takashi Kawamura, Hirohisa Takano, Kan Hiroshi Suzuki

## Abstract

This study examined how social isolation and subjective loneliness impacted subjective well-being across Japan, with the aim of offering policy recommendations. We analyzed data (*N* = 5,234, age:15-89) obtained through a Japanese population-based national survey conducted in March 2021. First, we found that mean life satisfaction was statistically lowest among the subgroup of respondents who lived alone (approximately 20% of the total), with more pronounced impacts in males. We also found that the frequency of social networking site (SNS) usage was significantly associated with overall life satisfaction, exhibiting a J-curve (three to four times per week was better than everyday usage). The frequency of interaction with others, number of SNS friends, employment/wage satisfaction, family budget/asset satisfaction, subjective health satisfaction, and subjective connectedness were also related to well-being in a dose-dependent manner (Trend P<0.001). Second, our multiple logistic regression model showed that the above relationships were independently and statistically significant (p<0.05). Moreover, a stratified analysis showed that living alone significantly affected these relationships, which suggests that this condition should specifically be addressed in the context of efforts aimed at enhancing subjective well-being. In addition, respondents who lived alone had low subjective connectedness, which also independently influenced well-being, as shown by our multivariate logistic model. In sum, these findings indicate that national well-being enhancement policies should particularly consider social isolation and loneliness, including issues stemming from low social connectedness.

## Introduction

Some countries were acknowledging issues of social isolation and loneliness as parts of their public health agendas even before the COVID-19 pandemic. For example, the United Kingdom appointed its first Minister for Loneliness in 2018; this was soon followed by efforts in Japan, where a Minister of Loneliness was appointed in 2021. Given that the proportions of people who live alone continually increase among older and working adults across the globe [1], loneliness has become one of the greatest public health challenges of our time. In this regard, the development and maintenance of meaningful social ties is thought to be essential for well-being [2]. By contrast, there is substantial evidence that the lack of social connections is associated with poor health [3]. A previous cohort study even found that loneliness was a significant mediator in the relationship between social network factors and depression [4].

Still, there are specific areas to consider when evaluating living alone as a status, since there are both positive and negative aspects to the arrangement. For instance, living alone can be challenging, but can also be more comfortable than living with others due to issues of convenience. Thus, there are conflicting findings in the literature, especially among earlier studies on the association between living alone and mental health. While some researchers have found that living alone does not constitute a risk factor for mental health [5,6], others have reported contradictory evidence [7,8]. One prospective study showed that participants who lived alone had a 1.81-fold (CI = 1.46-2.23) higher purchase rate of antidepressants during the follow-up period than those who did not live alone [7], while another found that persons who lived alone (vs. married) were around twice as likely to have anxiety or depressive disorders [8]. While one cohort study found that loneliness predicted subsequent increases in depressive symptoms, but not vice versa [9], another reported that increases in depressive symptoms did predict loneliness [10]. Apart from mental health, living alone is also consistently associated with increased cancer incidence [11]. Finally, recent research has shown that the social network has an independent effect on the course of depression [12], which is an important factor to consider when examining the effects of social networking sites (SNS).

In sum, there is a continual need for research not only to clarify the relationship between living alone and well-being, but also to examine the modifying effects. Indeed, despite the large number of individuals who now live by themselves, there is only limited evidence on how SNS factors influence the connection between well-being and living alone. Given the need to target the issue of loneliness through evidence based policy making (EBPM), this study examined national cross-sectional data from Japanese residents aged 15 to 89 years to elucidate the factors that are related to well-being, with a particular focus on the modifying effects of living alone.

## Materials and Methods

### Study sample

We examined cross-sectional data collected through a Japanese nationwide internet survey that was conducted as part of a research project titled “Well-being Survey and Quality of life,” as led by the Cabinet Office (CAO) of the Japanese government since October 2019. We selected the most recent cross-sectional data (*N* = 5,234; age range of 15-89 years), which was collected in March 2021. The survey was conducted in accordance with the Statistics Law in Japan, which governs the statistical, legal, ethical, and other rules for surveys conducted by the government. Informed consent was obtained from all respondents. Since we obtained the data with permission from the CAO in concordance with all guidelines, ethics approval was not required for this study.

The survey questionnaires were distributed to the registrants of an Internet survey company, with data thus collected from more than 5,000 respondents. In this regard, it should be noted that residents living in the metropolitan areas were underweighted compared to the actual population. The dataset is publicly available from the CAO website: https://form.cao.go.jp/keizai2/opinion-0011.html

### Measures

As individual-level covariates, we considered gender, age (five-year increments), and whether respondents lived alone (vs. not). Here, living alone was defined as a household number of 1. To gauge subjective self-rated health, the survey asked participants to answer the following question using a 10-point Likert scale (0 = not satisfied at all, 10 = highly satisfied): “In general, how satisfied are you with your life?” To gauge subjective social connectedness, subjective satisfaction about employment/wage, subjective satisfaction about the family budget/asset, the survey asked participants to rate each item using the same 10-point Likert scale. We also constructed a continuous variable for life satisfaction (higher levels represent more satisfaction) as a main outcome variable. Finally, the survey asked participants to report on their frequency of interaction with others (i.e., nearly every day, three or four times per week, once per week, two or three times per month, sometimes over the year, once per year, and no one to interact with).

### Analytic strategy

For the descriptive analysis, we computed the means and standard deviations for all study variables. We then separately computed each of these for respondents who lived alone and those who did not live alone; the differences were compared by t-test after checking the bell curve distribution, assuming that living status may both directly influence life satisfaction and modify how other factors affect it.

For the multiple logistic regression analysis, we constructed a binary variable indicating whether general satisfaction exceeded the mean score (5.79 on a 10-point Likert scale); this was set as the objective variable. As for the explanatory variables, we used gender (female=0, male=1), age (five-years increments), living alone (yes=0, no=1), employment and wage satisfaction (10-point Likert scale), family budget and asset satisfaction (10-point Likert scale), subjective health satisfaction (10-point Likert scale), subjective connectedness (10-point Likert scale), and frequency of SNS use (8-point Likert scale, everyday as standard). This multivariate analysis was conducted to evaluate the effects of living alone by controlling for the covariates.

As an additional analysis, we also divided participants into the three following interaction frequency groups: interacting with others each week, each month, and rarely (including no interaction). To further investigate SNS status, we divided participants into the four following groups based on the number of listed friends: no SNS friends, 1-29 SNS friends, 30-99 SNS friends, and 100 or more SNS friends. Stratified graphic representations were also illustrated in figures. All statistical analyses were conducted using STATA 17.0 (StataCop, College Station, TX, USA) and R ver. 4.1.2 (R Foundation for Statistical Computing, Vienna, Austria).

## Results

Table 1 summarizes the key participant characteristics. As shown, the mean age was 44.1 ± 16.9 (n=5,234), with 48.6% males. The table also stratifies the data based on living status.

**Table 1.** Participant sociodemographic characteristics, including differences between those living alone and those not living alone

| Characteristics: | Overall | Total ($n=5,234$) | | Difference<br>p-value |
| --- | --- | --- | --- | --- |
| | | Living Alone ($n = 919$) | Not Alone ($n = 4,315$) | |
| Age, mean ± SD | $44.1 \pm 16.9$ | $40.4 \pm 17.1$ | $44.8 \pm 16.8$ | <0.0001 ** |
| Gender, Male, (%) | 48.6 | 51.9 | 47.9 | 0.021 * |
| Living status (alone), (%) | 17.6 | 100 | 0 | <0.0001 ** |
| Frequency of SNS use, mean ± SD | $3.02 \pm 2.81$ | $3.03 \pm 2.81$ | $3.02 \pm 2.82$ | 0.450 |
| Satisfaction with employment and wage, mean ± SD | $5.62 \pm 2.21$ | $5.37 \pm 2.26$ | $5.68 \pm 2.20$ | 0.0002 ** |
| Satisfaction with family budget and asset, mean ± SD | $5.73 \pm 2.34$ | $5.42 \pm 2.44$ | $5.80 \pm 2.31$ | <0.0001 ** |
| Subjective self-rated health, mean ± SD | $6.52 \pm 2.17$ | $6.16 \pm 2.22$ | $6.59 \pm 2.16$ | <0.0001 ** |
| Subjective social connectedness, mean ± SD | $6.33 \pm 2.02$ | $6.03 \pm 2.11$ | $6.39 \pm 1.99$ | <0.0001 ** |
| Frequency of real interactions with others, mean ± SD | $5.04 \pm 2.05$ | $4.70 \pm 2.18$ | $5.11 \pm 2.02$ | <0.0001 ** |
| General satisfaction, mean ± SD | $5.75 \pm 2.33$ | $5.19 \pm 2.42$ | $5.86 \pm 2.30$ | <0.0001 ** |

SD = standard deviation, SNS = social networking sites
Employment & Wage Satisfaction, Family Budget & Asset Satisfaction, Subjective Health Satisfaction, and Subjective Connectedness were rated on a 10-point scale.
Frequency of SNS Use was rated on an 8-point scale.
\* $P < 0.05$ ; \*\* $P < 0.001$

First, we found that mean general life satisfaction was significantly lower among participants who lived alone when compared to others (5.19 ± 2.42 vs 5.86 ± 2.30, p<0.0001), representing nearly one-fifth of the total sample (17.6% of 5,234) (Fig 1).

**Fig 1.**
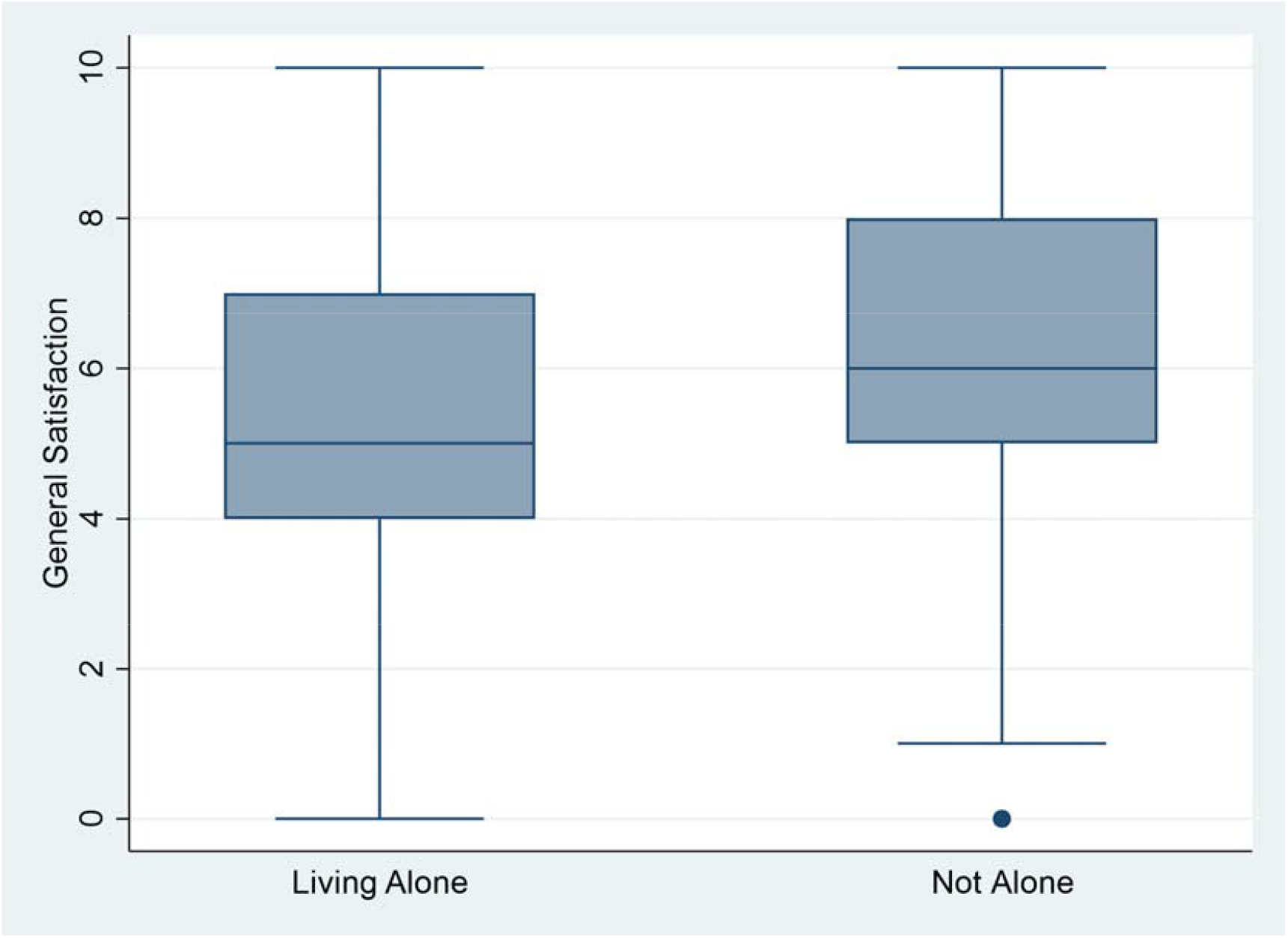
General satisfaction among participants who lived alone and others

Meanwhile, the mean age of participants who lived alone was significantly lower than that of those who did not (40.4 ± 17.1 vs 44.8 ± 16.8, p<0.0001). The percentage of males was also significantly higher among those who lived alone (51.9% vs 47.9%, p=0.021). The mean frequency of SNS use was about once per week (3 on the 8-point scale), with no significant differences between participants who lived alone and those who did not (3.03 ± 2.81 vs 3.02 ± 2.82, p=0.450). Satisfaction with employment and wage was significantly lower among participants who lived alone when compared to those who did not (5.37 ± 2.26 vs 5.68 ± 2.20, p=0.0002), as was satisfaction with the family budget and asset (5.42 ± 2.44 vs 5.80 ± 2.31, p<0.0001). Similarly, both subjective self-rated health (6.16 ± 2.22 vs 6.59 ± 2.16, p<0.0001) and subjective social connectedness (6.03 ± 2.11 vs 6.39 ± 1.99, p<0.0001) were significantly lower among participants who lived alone group when compared to those who did not.

As an important objective variable, we also considered the frequency of real-world interactions with others; here, the mean frequency was about once per month (5 on the 8-point scale), but this was lower among participants who lived alone when compared to those who did not (4.70 ± 2.18 vs 5.11 ± 2.02, p<0.0001). In addition, those who lived alone had lower satisfaction levels, including for general satisfaction. These findings reinforce the idea that national health and well-being policies should target individuals who live alone.

Next, we conducted a multiple logistic regression analysis among the various factors to determine whether general satisfaction exceeded the mean score (5.79 on the 10-point scale) (Table 2). Some variables were omitted from the logistic model due to multiple co-linearity.

**Table 2.** Multiple logistic regression analysis on factors influencing whether general satisfaction exceeded the mean score (5.79 on a 10-point scale)

|  | Odds Ratio [95% CI] | P value |
| --- | --- | --- |
| **Gender (female=0, male=1)** | 0.772 [0.660-0.903] | <0.001** |
| **Age (5-year increments)** | 1.034 [1.010-1.060] | 0.007** |
| **Living Alone (yes=0, no=1)** | 1.308 [1.063-1.611] | 0.011* |
| **Employment & Wage Satisfaction** | 1.124 [1.063-1.188] | <0.001** |
| **Family Budget & Asset Satisfaction** | 1.527 [1.446-1.613] | <0.001** |
| **Subjective Health Satisfaction** | 1.388 [1.321-1.459] | <0.001** |
| **Subjective Connectedness** | 1.326 [1.254-1.402] | <0.001** |
| **Frequency of SNS Use (everyday as standard)** | 0.813 [0.664-0.994] | <0.001** |
| **Constant** | 0.0015 | <0.001** |

SNS = social networking sites, CI = confidence interval
Employment & Wage Satisfaction, Family Budget & Asset Satisfaction, Subjective
Health Satisfaction, and Subjective Connectedness were rated on a 10-point scale.
Frequency of SNS Use was rated on an 8-point scale.
P for model <0.001 \*P<0.05; \*\*P<0.001

For each variable, the odds ratios (ORs) and 95% confidence intervals (CIs) showed significant and independent effects on general satisfaction. The OR and 95% CI for gender (female=0, male=1) was 0.772 [0.660-0.903], indicating that males (vs. females) more easily underran the mean satisfaction level after adjusting other variables (p<0.001). The OR and 95% CI for age (five-year increments) was 1.034 [1.010-1.060], indicating that aging was slightly more associated with easily surpassing the mean satisfaction level after adjusting for other variables (p=0.007). The ORs and 95% CIs for employment and wage satisfaction (10-point Likert scale) and family budget and asset satisfaction (10-point Likert scale) were 1.124 [1.063-1.188] and 1.527 [1.446-1.613], respectively, indicating that economic satisfaction helped participants surpass the mean general satisfaction level after adjusting for other variables (p<0.001, respectively). The OR and 95% CI for subjective health satisfaction (self-rated health on a 10-point Likert scale) was 1.388 [1.321-1.459], indicating a strong effect on general satisfaction after adjusting for other variables (p<0.001). Similarly, subjective connectedness (10-point Likert scale) had an OR and 95% CI of 1.326 [1.254-1.402], which also indicated a strong effect on general satisfaction after adjusting for other variables (p<0.001). The OR and 95% CI for SNS use frequency (8-point Likert scale, everyday as standard) was 0.772 [0.660-0.903], indicating that participants with less use more easily and significantly underran the mean satisfaction level after adjusting for other variables (p<0.001). Of particular note, the OR and 95% CI for living alone (yes=0, no=1) was 1.308 [1.063-1.611], indicating that participants who did not live not alone more easily and significantly surpassed the mean satisfaction level after adjusting for other variables (p=0.011).

Returning to the univariate analysis, we confirmed that the frequency of SNS usage was negatively associated with overall life satisfaction, showing a J-curve phenomenon (three to four times per week was better than everyday use) (Fig 2). The frequency of SNS use was nearly related to general satisfaction in a dose-dependent manner (Trend P < 0.001).

**Fig 2.**
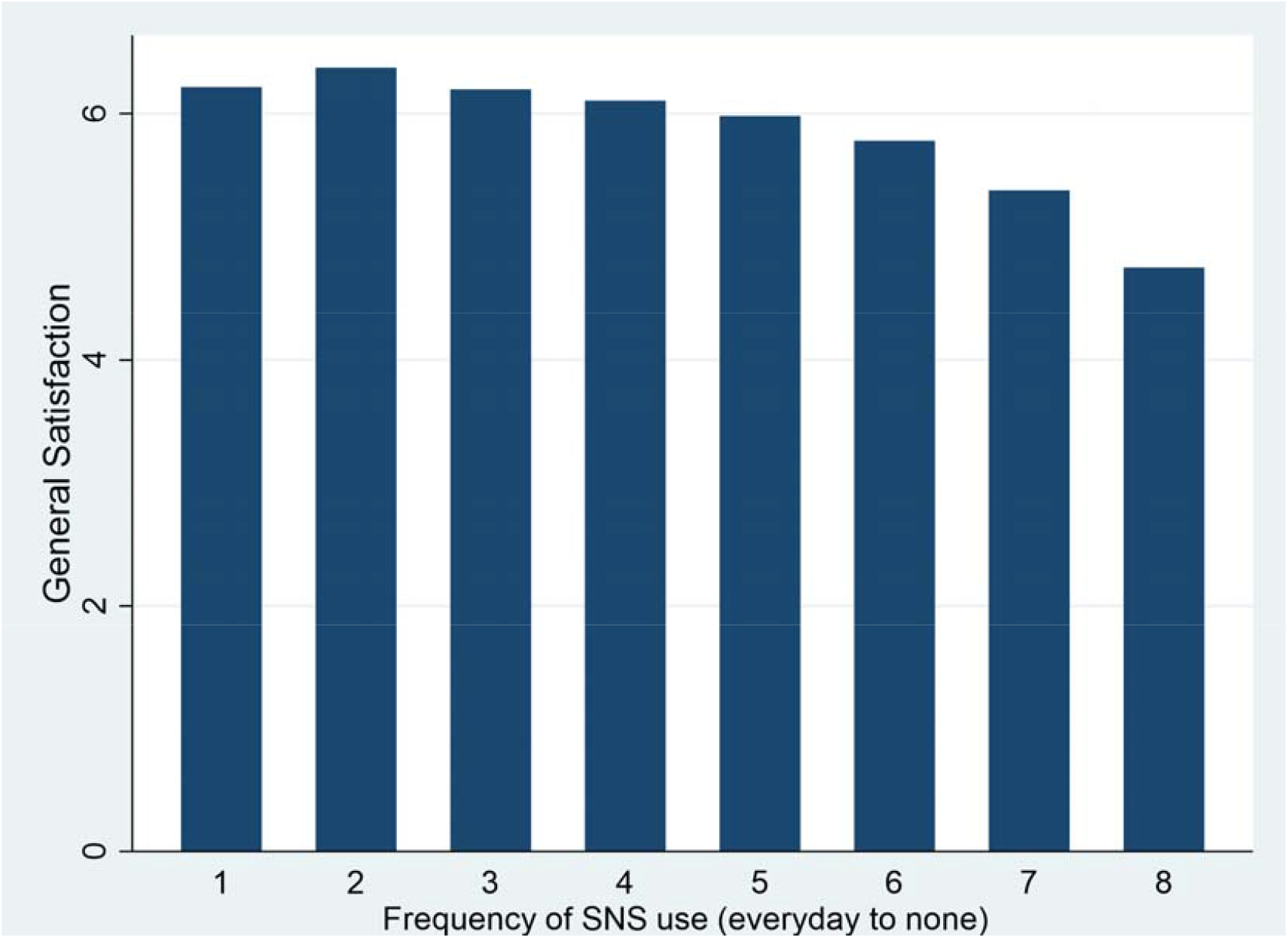
Frequency of SNS use and general satisfactions, showing a J-curve

As shown in Figs 3 through 6, employment/wage satisfaction, family budget/asset satisfaction, subjective health satisfaction, and subjective connectedness were each related to general satisfaction in a dose-dependent manner (Trend P < 0.001, respectively).

**Fig 3.**
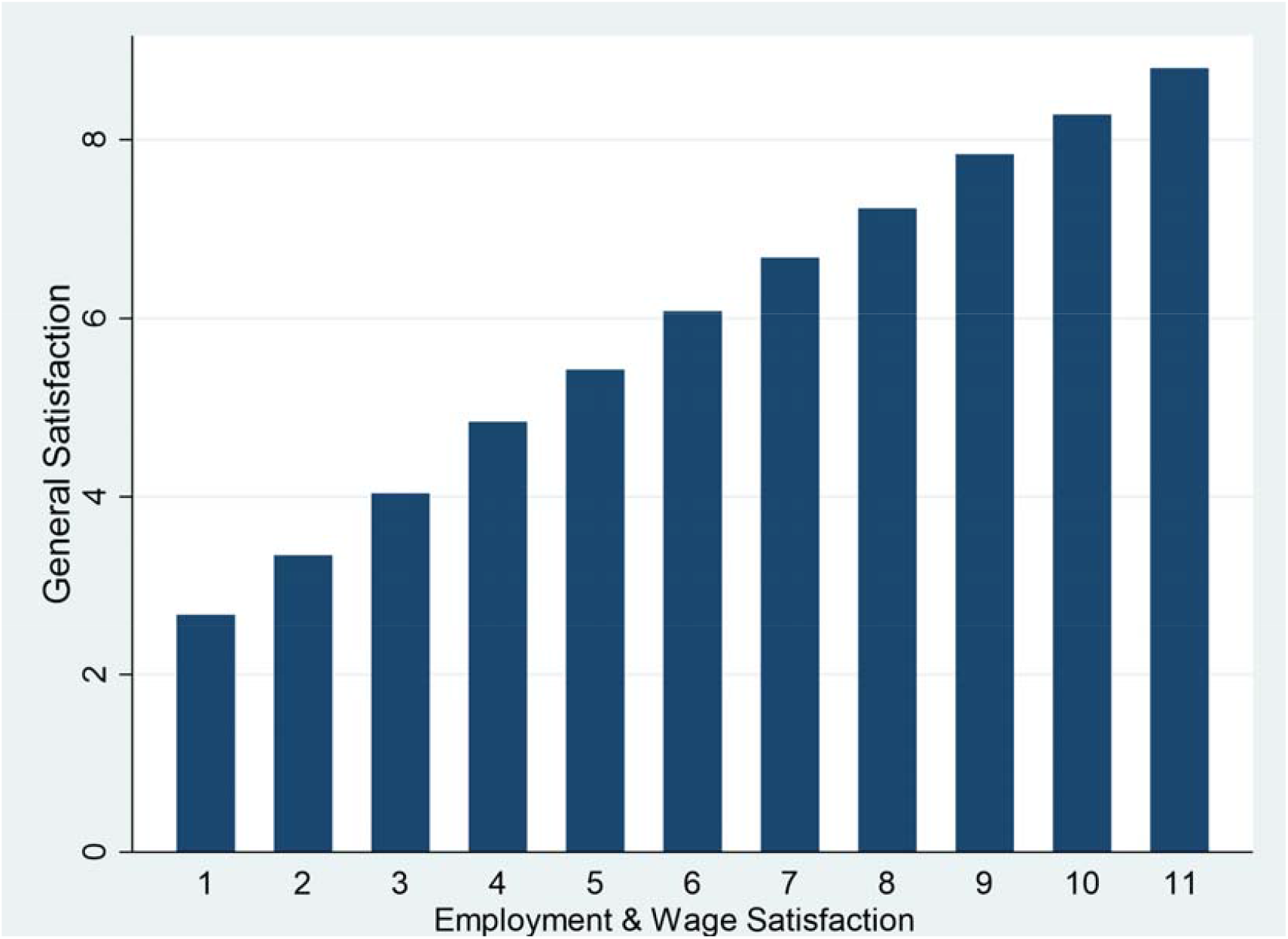
Satisfaction with employment & wage and general satisfaction

In regard to the J-curve phenomenon described above, we also conducted a stratified analysis. The phenomenon and overall dose-dependent manner were both observed in participants who lived alone, but not in those who did not live alone. Interestingly, the relationship between SNS use frequency and general satisfaction was more pronounced among participants who lived alone (Fig 7), with low or no SNS use showing remarkably low satisfaction in this group. On the other hand, although mean satisfaction was lower among participants who lived alone, frequent SNS use was associated with the same satisfaction among participants who lived alone and those who did not. It is remarkable that high and low SNS use showed anomalous results among those who lived alone.

**Fig 4.**
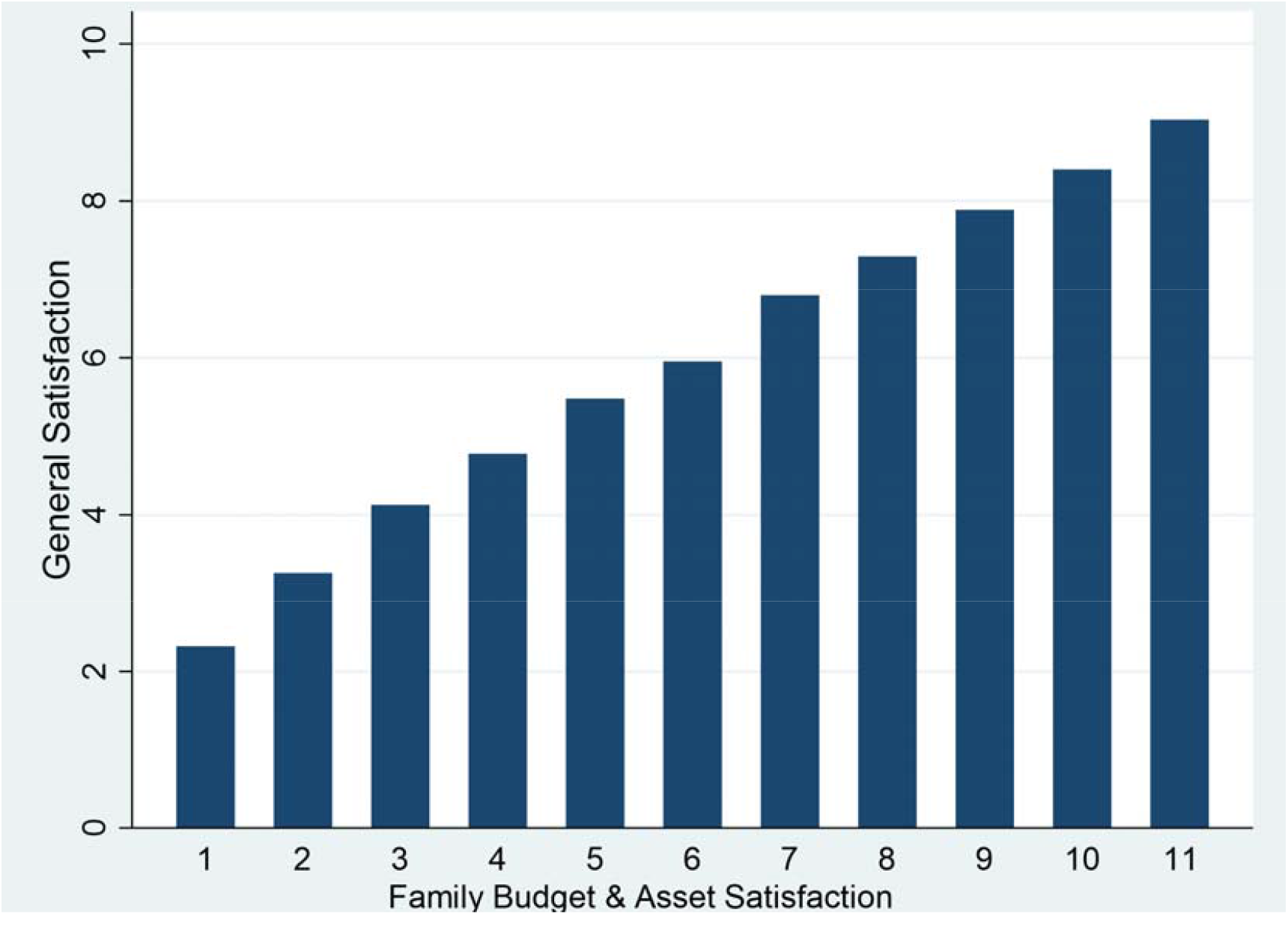
Satisfaction with the family budget & asset and general satisfaction

**Fig 5.**
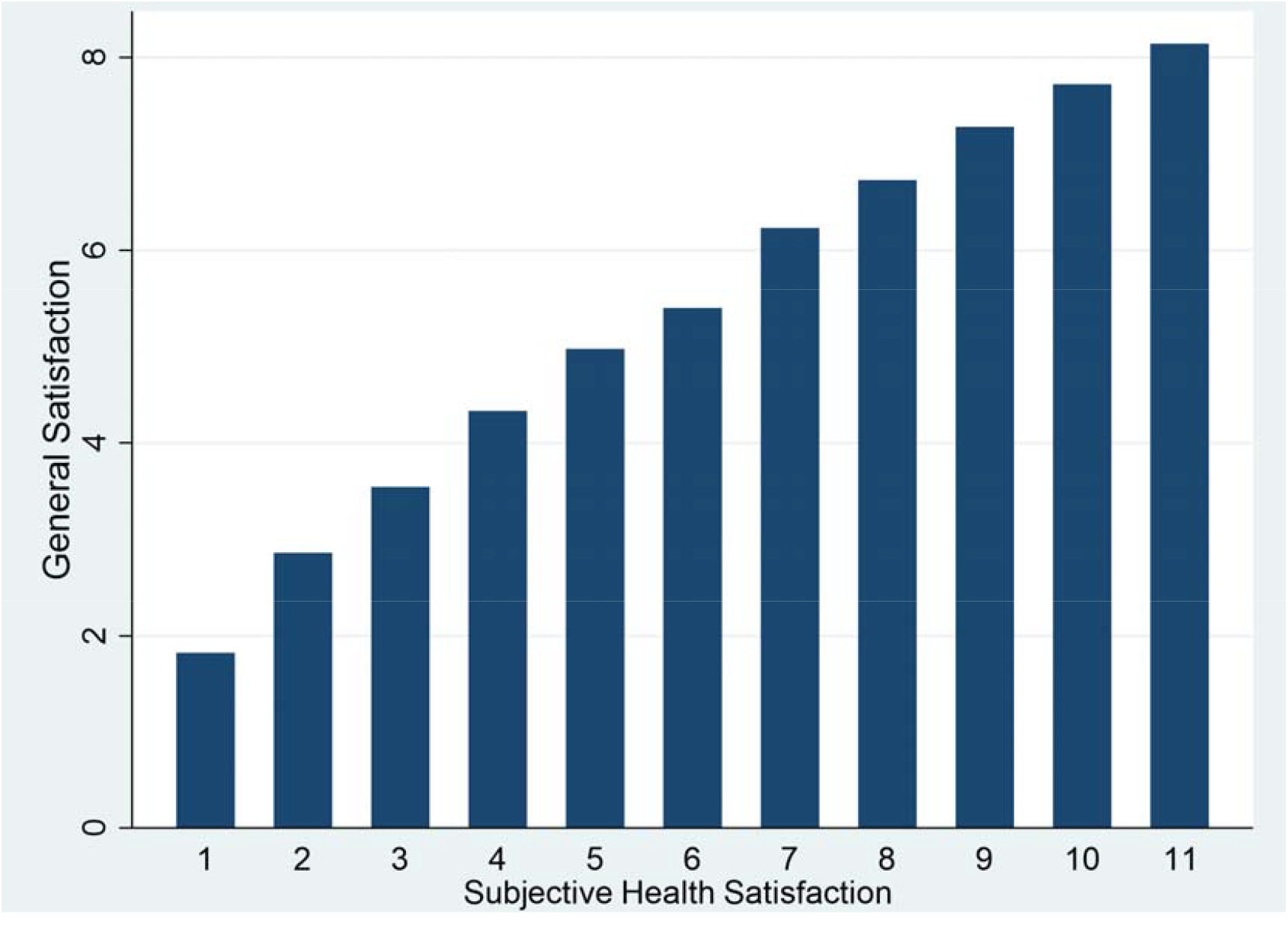
Subjective self-rated health and general satisfaction

**Fig 6.**
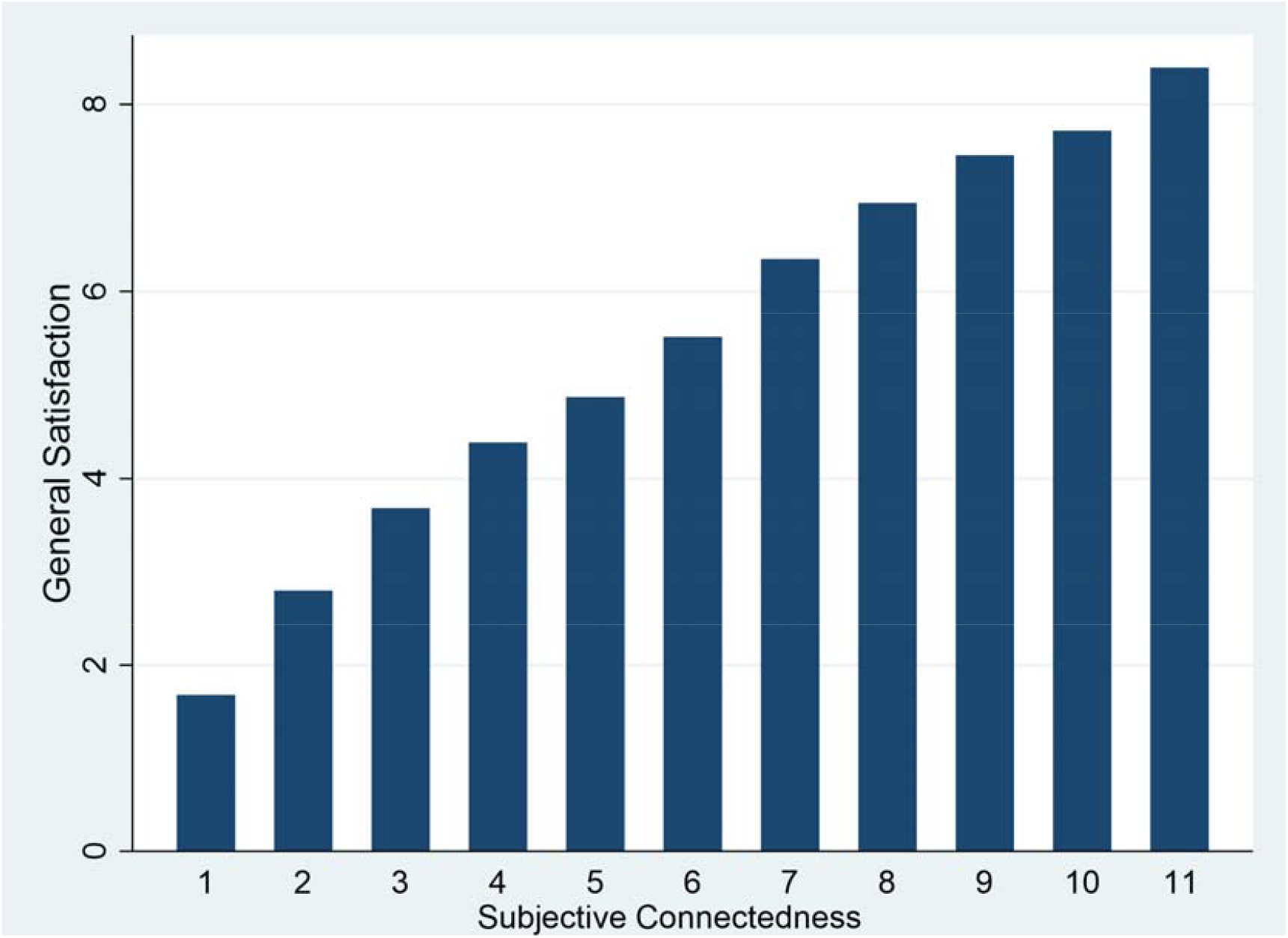
Subjective social connectedness and general satisfaction

**Fig 7.**
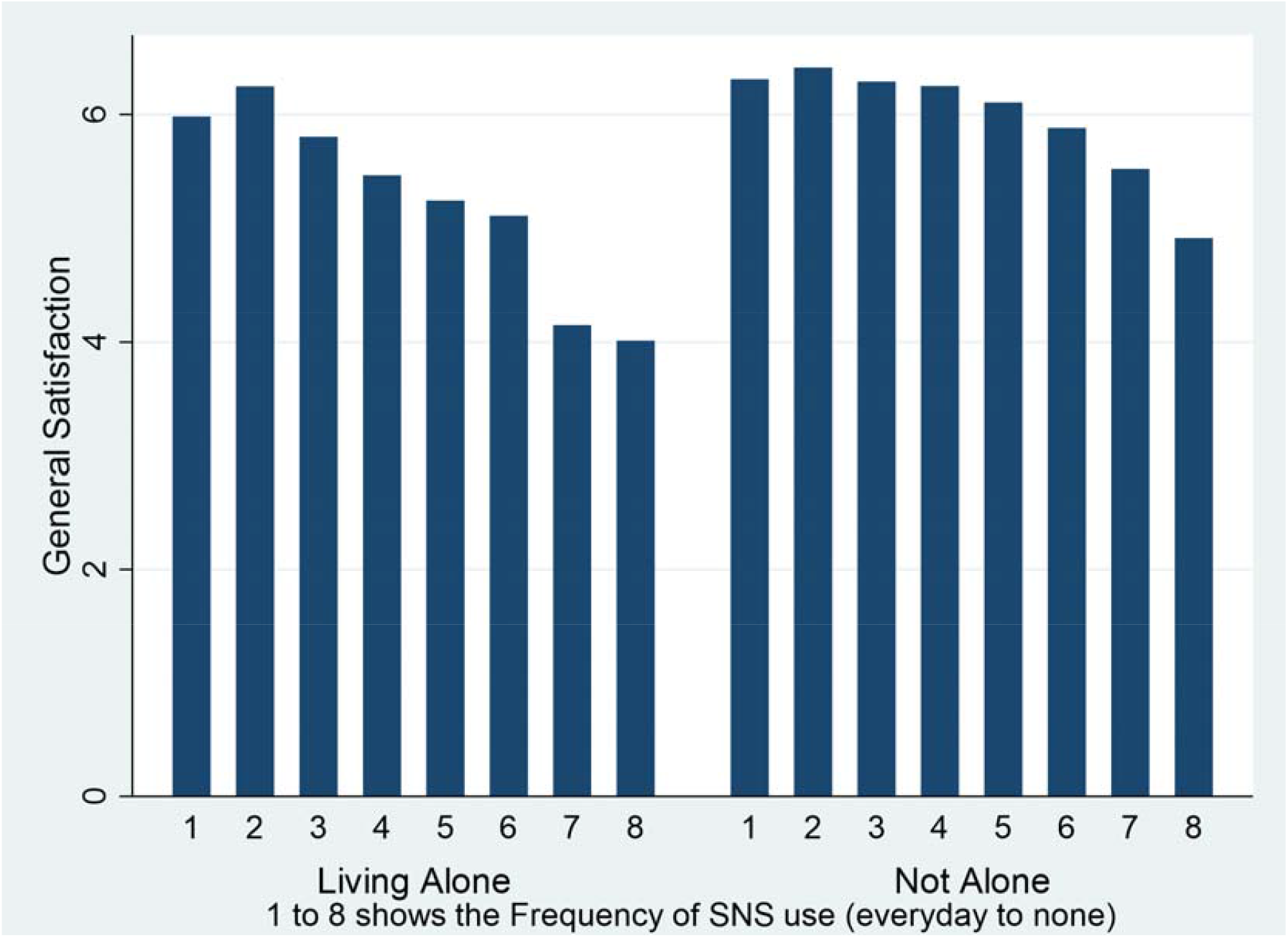
Frequency of SNS use and general satisfaction among participants who lived alone and others

As for gender, the difference in satisfaction levels between participants who lived alone and those who did not was larger for males (4.89 ± 2.50 vs 5.79 ± 2.31, p<0.0001) than for females (5.51 ± 2.28 vs 5.93 ± 2.29, p=0.0008) (Fig 8).

**Fig 8.**
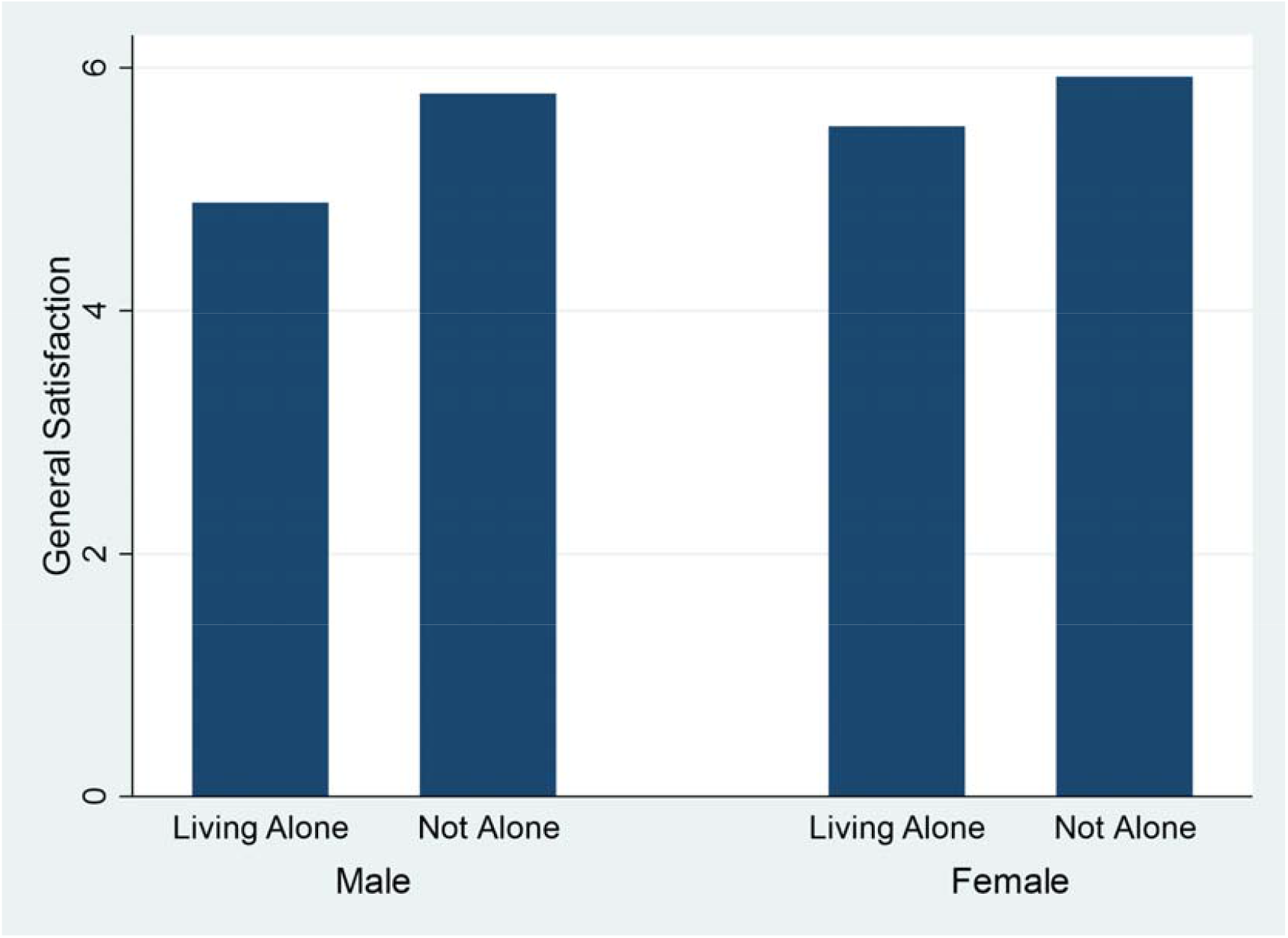
General satisfaction among participants who lived alone and others according to gender

Living alone also affected dose-dependent relations between subjective social connectedness and general satisfaction (Fig 9). In the low subjective connectedness group (especially those with 2 or 3 on the 1-11 point scale), general satisfaction seemed strongly suppressed, particularly for participants who lived alone. To the contrary, the high subjective connectedness group showed similar general satisfaction levels regardless of living status (Fig 9).

**Fig 9.**
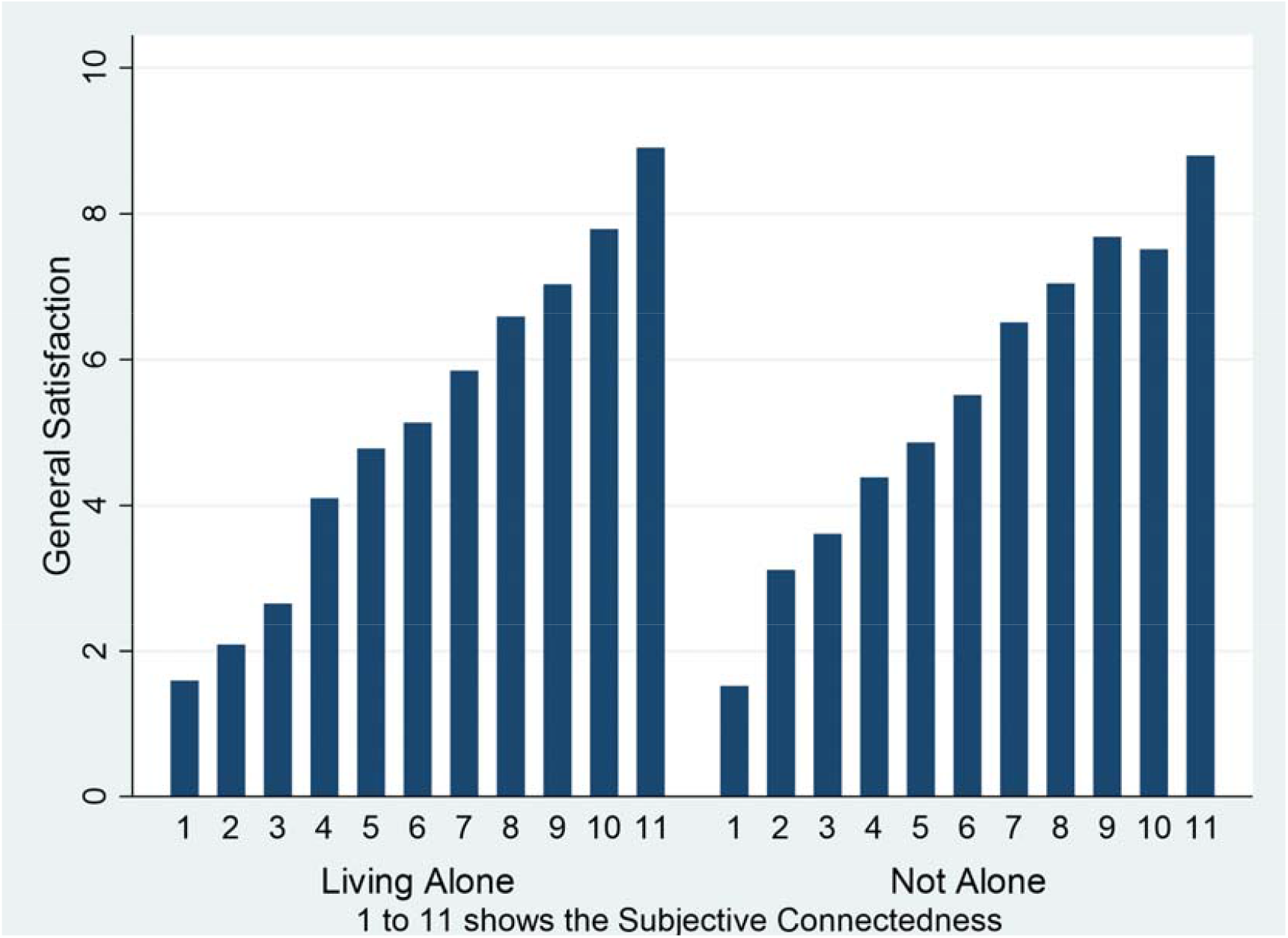
Subjective social connectedness and general satisfaction among participants who lived alone and others

We conducted another stratified analysis to investigate the modification effects on living alone. While the frequency of real interactions with others was related to general satisfaction for participants who lived alone and those who did, this relationship was more pronounced among those who lived alone (Fig 10). The frequent interaction group (weekly interactions) showed almost no differences in satisfaction levels based on living status.

**Fig 10.**
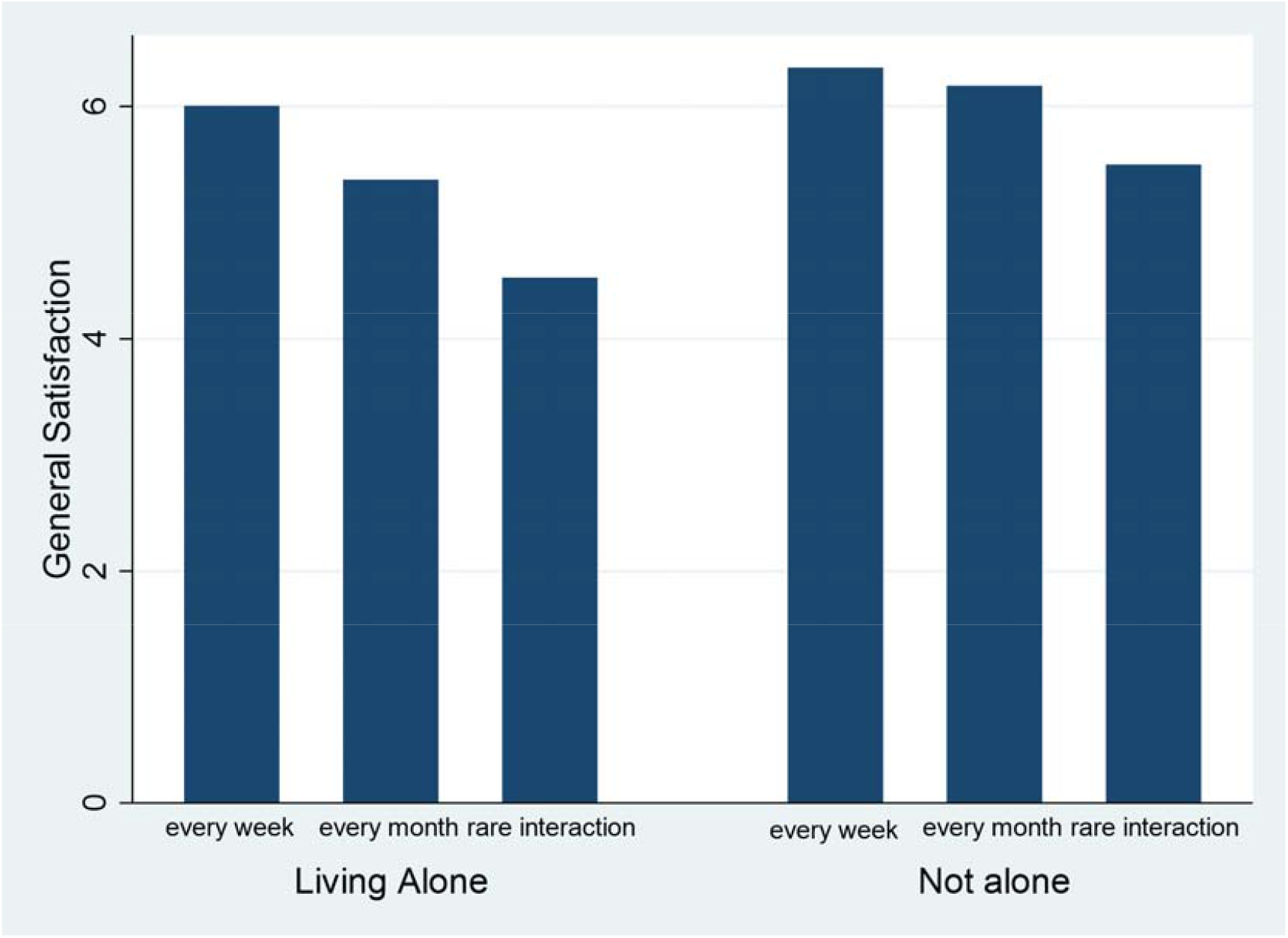
Frequency of interaction with others and general satisfaction among participants who lived alone and others

Similarly, the number of SNS friends was related to general satisfaction among participants who lived alone and those who did not, but this relationship was more pronounced among those who lived alone (Fig 11). Participants with fewer friends (less than 30) showed strong levels of low satisfaction, especially among those who lived alone.

**Fig 11.**
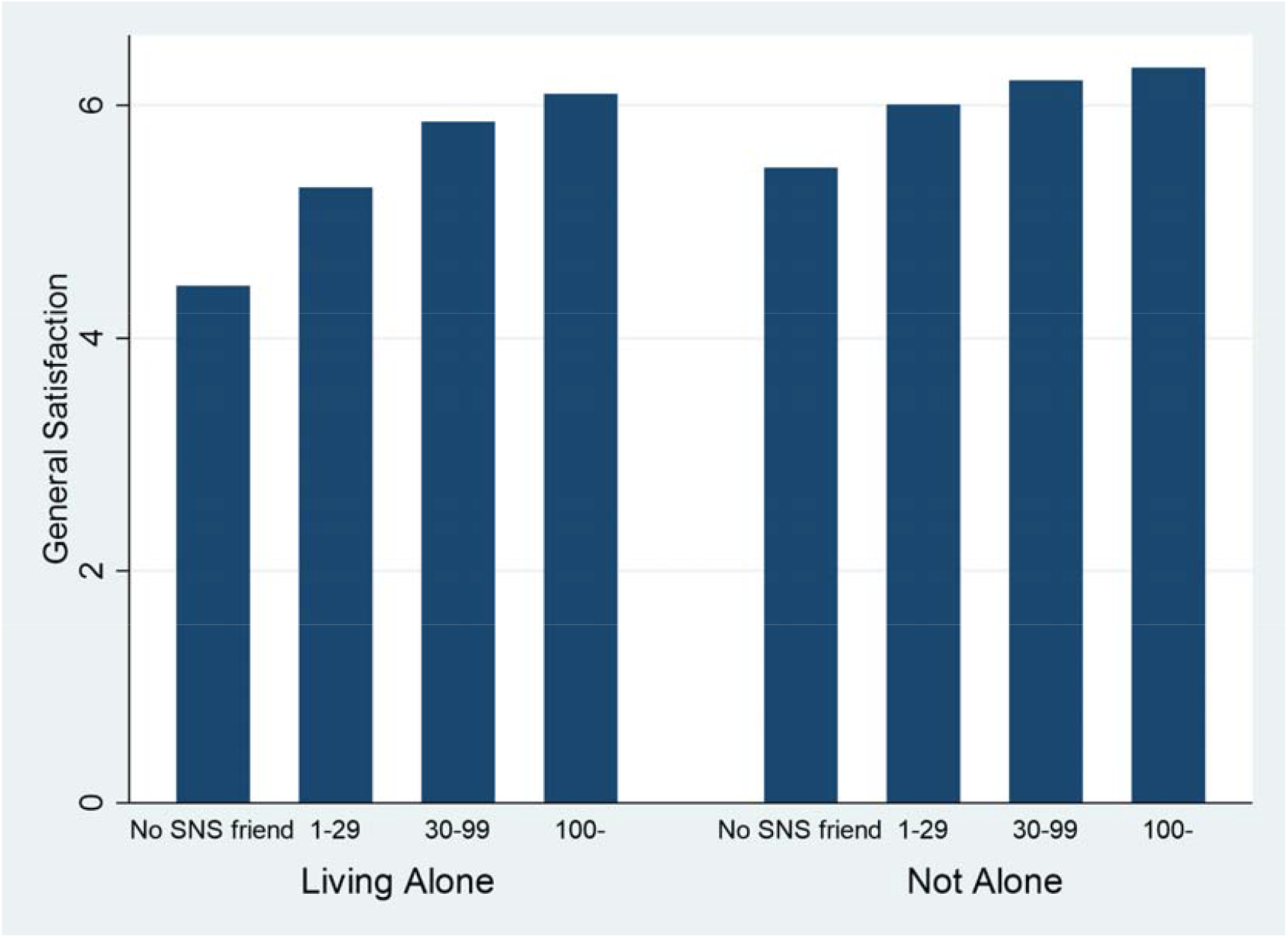
Number of SNS friends and general satisfaction among participants who lived alone and others

It is remarkable that participants who lived alone showed anomalous results in four areas (see Figs 7, 9, 10, and 11). The univariate and multivariate analyses consistently showed low general satisfaction among participants who lived alone, with independent factor effects. Living alone also influenced other relationships with satisfaction.

## Discussion

This study found that mean life satisfaction was statistically lower among individuals who lived alone, as compared with those in other living arrangements. This is consistent with previous reports [13–17], and thus largely expected, as individuals who live alone are often more vulnerable to unhappiness than those who live with their families or other partners. These findings highlight an important public health concern in Japan, where census data show an increasing number of single-person households among men and women aged 20 to 39 years, which is primarily the result of changes in marriage behaviors [18]. In this regard, a modern trend toward later marriage is expected to continue in many developed countries around the world. Since young unmarried Japanese individuals are now more likely to cohabitate with their parents when compared to counterparts in other wealthy countries, this issue may become especially pronounced in Japan. To address the increased potential for loneliness and other related issues, we suggest the need for interventions targeted at this population.

We also found that the frequency of SNS usage was significantly associated with overall life satisfaction, exhibiting a J-curve phenomenon (three to four times per week was better than everyday use). In other recent research, higher SNS consumption showed a negative correlation with happiness levels in Europe [19]. However, this does not directly contradict our current findings, as Japanese SNS consumption patterns tend to differ from those seen in other countries [20]. Moreover, we did find that everyday use (typical heavy use) was worse for well-being than more moderate use.

Next, we found that the frequency of interaction with others, number of SNS friends, employment/wage satisfaction, family budget/asset satisfaction, subjective health satisfaction, and subjective connectedness were related to well-being, each in a dose-dependent manner (Trend P<0.001). These relationships were both expected and consistent with previous findings [21–28]. Interestingly, most of these variables were statistically significant not only in the univariate model, but also in the multivariate model (Table 2). Our multiple logistic regression model showed statistically significant and independent relationships with several variables (p<0.05), including gender, age, living alone (or not), employment/wage satisfaction, family budget/asset satisfaction, subjective health satisfaction, subjective connectedness, and the frequency of SNS use.

Looking at effect strength, family budget/asset satisfaction imposed the largest effect, with an OR of 1.527. In this context, one increment of financial satisfaction on the 10-point scale increased the likelihood that general satisfaction would be above the mean by more than 50%. Other relatively large ORs included those for subjective health satisfaction, subjective connectedness, and living alone (or not), at 1.388, 1.326, and 1.308, respectively. Of particular note, living alone was a potent independent factor in lowering general satisfaction levels, even after controlling for other variables. By the same token, there is biological evidence that living alone is negatively related to gray matter volume through lowered subjective happiness [29]. This emphasizes the need to focus on the fact that individuals in Japan are increasingly living alone [18], including a trend toward later marriage, which is also occurring in many other developed countries.

In this study, living alone was associated with lower general satisfaction independently of other factors. Moreover, a stratified analysis (see Figs 7, 9, 10, and 11) showed that living alone influenced relationships between other factors and satisfaction. This underscores the major impacts that living alone has by itself. For example, the relationship between SNS use frequency and general satisfaction was more pronounced in participants who lived alone when compared to those who did not (Fig 7). For those who lived alone, low or no SNS use was associated with remarkably low satisfaction, while frequent SNS use was associated with the same satisfaction levels found among participants who did not live alone. Looking at participants with low subjective connectedness (especially those with a 2 or 3 on the 1-11 point scale), general satisfaction was strongly suppressed, especially among those who lived alone. These individuals represent good intervention candidates, as higher levels of subjective connectedness were associated with similar boosts in general satisfaction among participants who lived alone and those who did not; that is, satisfaction increased at similar rates with higher levels of connectedness regardless of living status (Fig 9).

While the frequency of real interaction with others was also related to general satisfaction across the sample, this was more pronounced among participants who lived alone (Fig 10). Since the frequent interaction group (weekly real interactions) showed highly similar levels of satisfaction regardless of living status, interventions may achieve substantial benefits by encouraging more interaction, especially among individuals who live alone.

Similarly, the number of SNS friends was related to general satisfaction across the sample, but this was also more pronounced among participants who lived alone (Fig 11). Because a lower number of friends (less than 30) was especially associated with low satisfaction levels in participants who lived alone, these individuals may be good candidates for relevant interventions. In sum, these results, especially those showing that living alone affects other relationships, suggest the potential efficacy of practical interventions aimed at enhancing subjective well-being. Moreover, such interventions may achieve additional success if they are tailored based on specific individual factors. For example, men who lived alone were even more vulnerable to low well-being than women with the same status, as evidenced by significantly lower satisfaction levels in that subsample (Fig 8).

Finally, individuals who lived alone (an objective state) showed low subjective connectedness, which also had independent effects on well-being, as shown via the multivariate logistic model (Table 2). While social connectedness and loneliness are considered opposite ends of the same subjective continuum [30,31], social isolation and participation are considered to exist along a continuum of objective social contact [32]. Interventions should be developed in consideration of both continua. In this regard, a recent meta-analysis showed that providing adults with opportunities to interact with others led to significant increases in objective social contact, while psychological strategies aimed at distress management led to significant improvements in the quality of social connections [33]. Due to the critical importance of subjective and objective outcomes, we believe that health policies should not only consider measures that reflect living alone status (an objective state), but should also consider loneliness (subjectively low social connectedness).

### Limitations

Our findings are based on data from a large national sample, and thus elucidate important cross-sectional relationship between variables that are known to impact well-being. However, there were also some limitations, in addition to the potential for preexisting measurement errors and biases stemming from the self-administered nature of the questionnaires. First, the cross-sectional design prohibited us from establishing causality between variables. Second, we assessed several areas of satisfaction based on a 10-point Likert scale, which may affect the sensitivity and specificity of rated items, especially in different cultural settings (e.g., Japanese individuals generally tend to provide modest answers). Third, the study sample was recruited via the Internet. In this regard, government data show that over 80% of people in Japan use the Internet, with rates exceeding 90% among those aged 13 to 59 years, as of 2020 [34]. Thus, the recruitment method may have resulted in selection bias. Despite these limitations, this study makes important contributions due to its consistent findings that individuals who live alone are more vulnerable in terms of well-being, especially in Japan.

## Conclusions

This study examined a variety of factors related to well-being in Japan, with a particular focus on the modifying effects of living alone. Our stratified analysis showed that living alone significantly affected well-being both independently and through other factors, thus suggesting that health interventions should target this condition. In particular, public health policies should work to enhance well-being by promoting or supporting moderate SNS use among individuals who live alone, especially those who are male and report infrequent usage. To alleviate the negative impacts of social isolation and loneliness, policymakers should also focus on healthy lifestyle factors, including interactions with others, SNS friendships, and subjective social connectedness. Indeed, the current findings imply that both social isolation and loneliness (i.e., low social connectedness) independently affect general satisfaction. In addition to addressing living alone status itself (an objective state), it is also desirable to focus on subjective loneliness and low subjective connectedness, especially considering evidence that individuals with limited objective social contact are likely to benefit from increased interaction frequency, including interactions that occur in the group context [35,36]. Finally, psychological strategies that focus on cognitive/behavioral processes and target maladaptive factors have also shown effectiveness [37].

Both the increasing rate at which individuals are living alone and issues that arise thereof should be targeted through multidimensional EBPM approaches that consider the best available evidence. This points to the need for continued research.

## Data Availability

The dataset is publicly available from the Cabinet Office (CAO) of the Japanese government website: https://form.cao.go.jp/keizai2/opinion-0011.html

## Acknowledgements

Prof. Takashi Oshio (Institute of Economic Research, Hitotsubashi University) provided encouragement and useful advice on this study through personal communications.

## Author Contributions

**Conceptualization:** Koichi Miyaki

**Data curation:** Koichi Miyaki

**Statistical analysis:** Koichi Miyaki

**Funding acquisition:** Hiroshi Kan Suzuki

**Investigation:** Koichi Miyaki

**Methodology:** Koichi Miyaki

**Project administration:** Hirohisa Takano, Hiroshi Kan Suzuki

**Supervision:** Takashi Kawamura

**Writing – original draft:** Koichi Miyaki

**Writing – review & editing:** Takashi Kawamura, Hirohisa Takano, Hiroshi Kan Suzuki

## Funding

KM was supported by a grant from the Graduate School of Public Policy, University of Tokyo and the Research Institute of Occupational Mental Health (RIOMH). The funders had no role in the study design, data collection, analysis, decision to publish, or preparation of the manuscript.

## Conflict of interest

The authors declare no competing interests.

## Informed consent

Comprehensive informed consent was obtained from all participants. The survey was conducted in accordance with the Statistics Law in Japan, which governs the statistical, legal, ethical, and other rules for surveys conducted by the government. The ethical oversight was waived by the decision of Institutional Review Board of Research Institute of Occupational Mental Health.

## Notes

### Competing Interest Statement

The authors have declared no competing interest.

### Funding Statement

The authors received no specific funding for this work.

### Author Declarations

The ethical oversight was waived by the decision of Institutional Review Board of Research Institute of Occupational Mental Health.

## References

1. Jamieson L, Simpson R. Living alone. Globalization, identity and belonging. Hampshire: Palgrave Macmillan studies in family and intimate life. 2013.

2. Cacioppo JT, Patrick W. Loneliness: human nature and the need for social connection. New York: W W Norton & Co.; 2008. p. 317.

3. Holt-Lunstad J. Why social relationships are important for physical health: A systems approach to understanding and modifying risk and protection. Annu Rev Psychol. 2018;69: 437–458. doi: 10.1146/annurev-psych-122216-011902.

4. Santini ZI, Fiori KL, Feeney J, Tyrovolas S, Haro JM, Koyanagi A. Social relationships, loneliness, and mental health among older men and women in Ireland: a prospective community-based study. J Affect Disord. 2016;204: 59–69. doi: 10.1016/j.jad.2016.06.032.

5. Michael YL, Berkman LF, Colditz GA, Kawachi I. Living arrangements, social integration, and change in functional health status. Am J Epidemiol. 2001;153: 123–131. doi: 10.1093/aje/153.2.123.

6. Kawamoto R, Yoshida O, Oka Y, Kodama A. Influence of living alone on emotional well-being in community-dwelling elderly persons. Geriatr Gerontol Int. 2005;5: 152–158. doi: 10.1111/j.1447-0594.2005.00285.x.

7. Pulkki-Råback L, Kivimäki M, Ahola K, Joutsenniemi K, Elovainio M, Rossi H, et al. Living alone and antidepressant medication use: a prospective study in a working-age population. BMC Public Health. 2012;12: 236. doi: 10.1186/1471-2458-12-236.

8. Joutsenniemi K, Martelin T, Martikainen P, Pirkola S, Koskinen S. Living arrangements and mental health in Finland. J Epidemiol Community Health. 2006;60: 468–475. doi: 10.1136/jech.2005.040741.

9. Cacioppo JT, Hawkley LC, Thisted RA. Perceived social isolation makes me sad: 5-year cross-lagged analyses of loneliness and depressive symptomatology in the Chicago Health, Aging, and Social Relations Study. Psychol Aging. 2010;25: 453–463. doi: 10.1037/a0017216.

10. Dahlberg L, Andersson L, McKee KJ, Lennartsson C. Predictors of loneliness among older women and men in Sweden: a national longitudinal study. Aging Ment Health. 2014;7863: 1–9.

11. Elovainio M, Lumme S, Arffman M, Manderbacka K, Pukkala E, Hakulinen C. Living alone as a risk factor for cancer incidence, case-fatality and all-cause mortality: A nationwide registry study. SSM Popul Health. 2021;15: 100826. doi: 10.1016/j.ssmph.2021.100826.

12. Houtjes W, Van Meijel B, Van De Ven PM, Deeg D, van Tilburg T, Beekman A. The impact of an unfavorable depression course on network size and loneliness in older people: a longitudinal study in the community. Int J Geriatr Psychiatry. 2014;29: 1010–1017. doi: 10.1002/gps.4091.

13. Hwang EJ, Sim IO. The structural equation modeling of personal aspects, environmental aspects, and happiness among older adults living alone: a cross-sectional study. BMC Geriatr. 2021;21: 479. doi: 10.1186/s12877-021-02430-4.

14. Noguchi T, Nojima I, Inoue-Hirakawa T, Sugiura H. Role of non-face-to-face social contacts in moderating the association between living alone and mental health among community-dwelling older adults: a cross-sectional study. Public Health. 2021;194: 25–28. doi: 10.1016/j.puhe.2021.02.016.

15. Hwang EJ, Sim IO. Association of living arrangements with happiness attributes among older adults. BMC Geriatr. 2021;21: 100. doi: 10.1186/s12877-021-02017-z, PMID: 33541268, PMCID: PMC7860621.

16. Lin YT, Chen M, Ho CC, Lee TS. Relationships among leisure physical activity, sedentary lifestyle, physical fitness, and happiness in adults 65 years or older in Taiwan. Int J Environ Res Public Health. 2020;17: 5235. doi: 10.3390/ijerph17145235.

17. Kim J, Song Y, Kim T, Park K. Predictors of happiness among older Korean women living alone. Geriatr Gerontol Int. 2019;19: 352–356. doi: 10.1111/ggi.13615.

18. Raymo JM. Living alone in Japan: relationships with happiness and health. Demogr Res. 2015;32: 1267–1298. doi: 10.4054/DemRes.2015.32.46.

19. Muñiz-Velázquez JA, Gómez-Baya D, Lozano Delmar J. Exploratory study of the relationship Between happiness and the rise of media consumption During COVID-19 confinement. Front Psychol. 2021;12: 566517. doi: 10.3389/fpsyg.2021.566517.

20. Sakurai R, Nemoto Y, Mastunaga H, Fujiwara Y. Who is mentally healthy? Mental health profiles of Japanese social networking service users with a focus on line, Facebook, Twitter, and Instagram. PLOS ONE. 2021;16: e0246090. doi: 10.1371/journal.pone.0246090.

21. Oshio T, Kimura H, Nishizaki T, Omori T. Association between the use of social networking sites, perceived social support, and life satisfaction: evidence from a population-based survey in Japan. PLOS ONE. 2020;15: e0244199. doi: 10.1371/journal.pone.0244199.

22. Jagtiani MR, Kelly Y, Fancourt D, Shelton N, Scholes S. #StateOfMind: family meal frequency moderates the association Between time on social networking sites and well-being Among U.K. Young adults. Cyberpsychol Behav Soc Netw. 2019;22: 753–760. doi: 10.1089/cyber.2019.0338.

23. Baek YM, Bae Y, Jang H. Social and parasocial relationships on social network sites and their differential relationships with users’ psychological well-being. Cyberpsychol Behav Soc Netw. 2013;16: 512–517. doi: 10.1089/cyber.2012.0510.

24. Platts K, Breckon J, Marshall E. Enforced home-working under lockdown and its impact on employee wellbeing: a cross-sectional study. BMC Public Health. 2022;22: 199. doi: 10.1186/s12889-022-12630-1.

25. Park S, Park C, Sung JH. How does the involuntary choice of self-employment affect subjective well-being in small-sized business workers? A cross-sectional study from the fifth Korean working conditions survey. Int J Environ Res Public Health. 2022;19: 1011. doi: 10.3390/ijerph19021011.

26. Martin SD, Urban RW, Johnson AH, Magner D, Wilson JE, Zhang Y. Health-related behaviors, self-rated health, and predictors of stress and well-being in nursing students. J Prof Nurs. 2022;38: 45–53. doi: 10.1016/j.profnurs.2021.11.008.

27. Badri MA, Alkhaili M, Aldhaheri H, Alnahyan H, Yang G, Albahar M, et al. Understanding the interactions of happiness, self-rated health, mental feelings, habit of eating healthy and sport/activities: A path model for Abu Dhabi. Nutrients. 2021;14: 55. doi: 10.3390/nu14010055.

28. Yuen EYN, Wilson CJ. The relationship between cancer caregiver burden and psychological outcomes: the moderating role of social connectedness. Curr Oncol. 2021;29: 14–26. doi: 10.3390/curroncol29010002.

29. Kokubun K, Pineda JCD, Yamakawa Y. Unhealthy lifestyles and brain condition: examining the relations of BMI, living alone, alcohol intake, short sleep, smoking, and lack of exercise with gray matter volume. PLOS ONE. 2021;16: e0255285. doi: 10.1371/journal.pone.0255285.

30. Fässberg MM, van Orden KA, Duberstein P, Erlangsen A, Lapierre S, Bodner E, et al. A systematic review of social factors and suicidal behavior in older adulthood. Int J Environ Res Public Health. 2012;9: 722–745. doi: 10.3390/ijerph9030722.

31. Kim H-J, Hong S, Kim M. Living arrangement, social connectedness, and life satisfaction among Korean older adults with physical disabilities: the results from the national survey on persons with disabilities. J Dev Phys Disabil. 2015;27: 307– 321. doi: 10.1007/s10882-014-9418-9.

32. Douglas H, Georgiou A, Westbrook J. Social participation as an indicator of successful aging: an overview of concepts and their associations with health. Aust Health Rev. 2017;41: 455–462. doi: 10.1071/AH16038.

33. Zagic D, Wuthrich VM, Rapee RM, Wolters N. Interventions to improve social connections: a systematic review and meta-analysis. Soc Psychiatry Psychiatr Epidemiol. 2021. doi: 10.1007/s00127-021-02191-w. Epub ahead of print. PMID: 34796368.

35. Haslam SA. Psychology in organizations: the social identity approach. London: SAGE; 2001.

36. Turner JC. Toward a cognitive redefinition of the social group. In: Tajfel H, editor Social identity and intergroup relations. Cambridge: Cambridge University Press; 1982. pp. 15–40.

37. Cacioppo JT, Hawkley LC. Perceived social isolation and cognition. Trends Cogn Sci. 2009;13: 447–454. doi: 10.1016/j.tics.2009.06.005.

34. Information & communications statistics database by Japanese Ministry of Internal Affairs and communications https://www.soumu.go.jp/johotsusintokei/statistics/statistics05.html.

